# Time-analysis of COVID-19 dispersion among health care workers and the general population

**DOI:** 10.1101/2021.08.07.21261433

**Authors:** Patricia A. F. Leme, Mehrsa Jalalizadeh, Franciele A. V. Dionato, Keini Buosi, Luciana S.B. Dal Col, Cristiane F. Giacomelli, Karen L. Ferrari, Ana Carolina Pagliarone, Lucas M. Gon, Cristiane L. Maia, Akbar A. Esfahani, Leonardo O. Reis

**Author notes:** Correspondence to: Leonardo Oliveira Reis, MD, MSc, PhD, UroScience, Pontifical Catholic University of Campinas (PUC-Campinas), R. John Boyd Dunlop, s/n, Campinas – São Paulo - Brasil - CEP: 13060-904.

## Abstract

**Introduction:** Heath care workers with direct (HCW-D) or indirect (HCW-A) patient contact represent 4.2% to 17.8% of COVID-19 cases. We evaluate the temporal COVID-19 infection behavior among HCW-D, HCW-A, and non-HCW.

**Methods:** From February 2020 to April 2021, trained nurses recorded age, gender, occupation, and symptoms in a COVID-19 testing outpatient health center. We allocated data into weekly time fractals and calculated the proportion of COVID-19 positive among HCW vs. non-HCW and incorporated an ARFIMA model (traditionally used in weather forecast) to predict future cases of COVID-19.

**Results:** Among 8,998 COVID-19 RT-PCR tests, 3,462 (42%) patients were HCW-D, and 933 (11%) were HCW-A. Overall, 1,914 (21.3%) returned positive, representing 27%, 25% and 19% of HCW-D, HCW-A and non-HCW, respectively. HCW-D or HCW-A were significantly more likely to test positive for COVID-19 than non-HCW (OR=1.5, p<0.0001). The percentage of positive to negative test results remained steady over time. In the positive cases, the percentage of HCW to non-HCW declined significantly over time (Mann-Kendal trend test: tau=-0.58, p<0.0001). Our ARFIMA model showed a long-memory infection pattern in the occurrence of new COVID-19 cases lasting for months. Average error was 1.9 cases per week comparing predicted to actual values three months later (May-July 2021).

**Conclusion:** HCW have a sustained 50% higher risk of COVID-19 positivity in the pandemic. Time-series analysis showed a long-memory infection pattern with virus spread mainly among HCWs before the general population. The tool http://wdchealth.covid-map.com/shiny/covid-map/ will be updated according to population previous infection and vaccination impact.

## INTRODUCTION

The SARS-CoV-2 drove worldwide attention since its outbreak in Wuhan, China, in 2019 [1]. The biggest concerns were about the spread velocity and the capacity of healthcare systems to give support to the population [2,3]. The healthcare workers (HCW) were much requested to support the increasing need for medical assistance while facing a new and highly contagious disease. Thus, there was a big concern about healthcare workers’ infections and mental health [4]. Reports from China, USA, and Italy showed that HCW represent 4.2%, 17.8%, and 9% of the infected population, respectively [5].

It is easy to assume that healthcare workers have a higher risk of infection than the general population. It comes from the frequent contact with infected people added to situations of increased risk, namely closed places, and procedures that increase the viral quantity in the air. On the other hand, individual protection devices and protection protocols like patient isolation help to limit that risk [6-8]. A prospective study based on smartphone application found a hazard ratio of 11.6 for HCW positive test [9], however, the increased risk might be subject to bias, such as the proportion at the studied population and the access for testing. A systematic review of records between November 2019 and May 2020 concluded that overall infection among HCW followed the general population [10]. However, this systemic review used information gathered in a very short amount of time, when the pandemic was only beginning and used low-quality data.

The COVID-19 proved to be a highly contagious disease. It took just three months for the World Health Organization to declare the SARS-CoV-2 pandemic [11]. The spread velocity and impact over healthcare systems were different among different countries. This study aims to bring evidence from one of the most important universities in Brazil during the pandemic. The high prevalence of healthcare workers made it possible to compare them to non-HCW and present the temporal risk of infection. Moreover, we propose a tool to predict the number of positive cases in the future using a hybrid forecast machine learning algorithm.

## METHODS

After local ethics committee approval number 4.173.069, all information was gathered from the University of Campinas outpatient health center, Campinas, São Paulo, one of the biggest universities in Brazil. All patients over 16 years old referred for COVID RT-PCR testing were added to the database. Trained nurses recorded each patient’s data: age, gender, occupation, address, and symptoms. All positive patients were followed up by phone for 14 days or until symptom-free.

We defined HCW as any occupation that involves direct (HCW-D) or indirect contact with patients (healthcare associates or HCW-A). We compared these two groups to those without healthcare-related jobs (non-HCW).

All statistical analyses were performed using R version 4.0.2 (2020-06-22) on RStudio platform version 1.3.1073 and using the following packages: tidyverse, lubridate, forecast, quantreg, splines, ggmap, pracma, fractaldim, and janitor. Normality for samples of n<5,000 was calculated using the Shapiro-Wilk test, and for n>5,000 the shape of the histogram was considered. The significance was considered when p<0.05.

### Time series analysis and forecast

Data was allocated into weekly time fractals. The proportion of female gender, positive results, and HCW were calculated per week and the Mann-Kendall trend test was used to detect change over time. To better understand outcomes and detect underlying relationships between the occurrence of positive cases in time, we transformed the count of positive COVID cases to fractal dimension via the R package fractaldim. This transformation reveals the self-similarity of the spread of infections across time and shows its memory process. After confirming a long memory process for COVID, we avoided traditionally used forecasting models, (such as AutoRegressive Integrated Moving Average [ARIMA] models or machine learning models) and instead, incorporated ARFIMA (AutoRegressive Fractionally Integrated Moving Average) forecasting model, which accounts for long memory by fractional differencing the time series. The ARFIMA model is like the Box-Jenkins ARIMA model, except that the integrated part of the ARIMA model can be a fractional number defined that is the inverse of the Hurst parameter [12]. The fractional differencing allows a better representation of the data since it decays slower than other types of time series.

We tested our models forecasting ability using data gathered 3 months later (May-July 2021) to estimate the reliability of our 12 week forecasting tool while keeping the investigators blind to actual values.

## RESULTS

From February 2020 to April 2021, a total of 8,998 COVID-19 RT-PCR tests were performed; a total of 1,914 (21.3%) of them returned positive, 70.6% returned negative, and 8% were either inconclusive or not available. Inconclusive or NA results were excluded for further analysis. Fifty patients needed hospitalization, and three died. The mean age of participants was 38.0, minimum 16 and maximum 95 years old. COVID-19 positive patients were on average 1.8 years older (95% CI: 1.2 - 2.4 years, p<0.0001). There was a gender discrepancy, with 70% of the patients being female.

### Occupation based analysis

A total of 3,462 (42%) patients were HCW-D, 933 (11%) HCW-A, and 3,877 (47%) patients were non-HCW. The gender discrepancy was significant in healthcare-related groups but not in the non-HCW: 82% of HCW-D and 81% of HCW-A were female, while only 57% of non-HCW were female (**Figure 1**). COVID-19 was confirmed in 27%, 25%, and 19% of HCW-D, HCW-A, and non-HCW, respectively. Probability of positive result did not depend significantly on gender (OR = 1.12, 95% CI 1.00 – 1.25, p = 0.041). Mean age was significantly lower in the non-HCW group: 35.6 ± 13.6 years versus 41.0 ±11.4 for HCW-D and 39.9 ±10.2 for HCW-A (p<0.0001, one-sided ANOVA, **Figure 2**).

**Figure 1.**
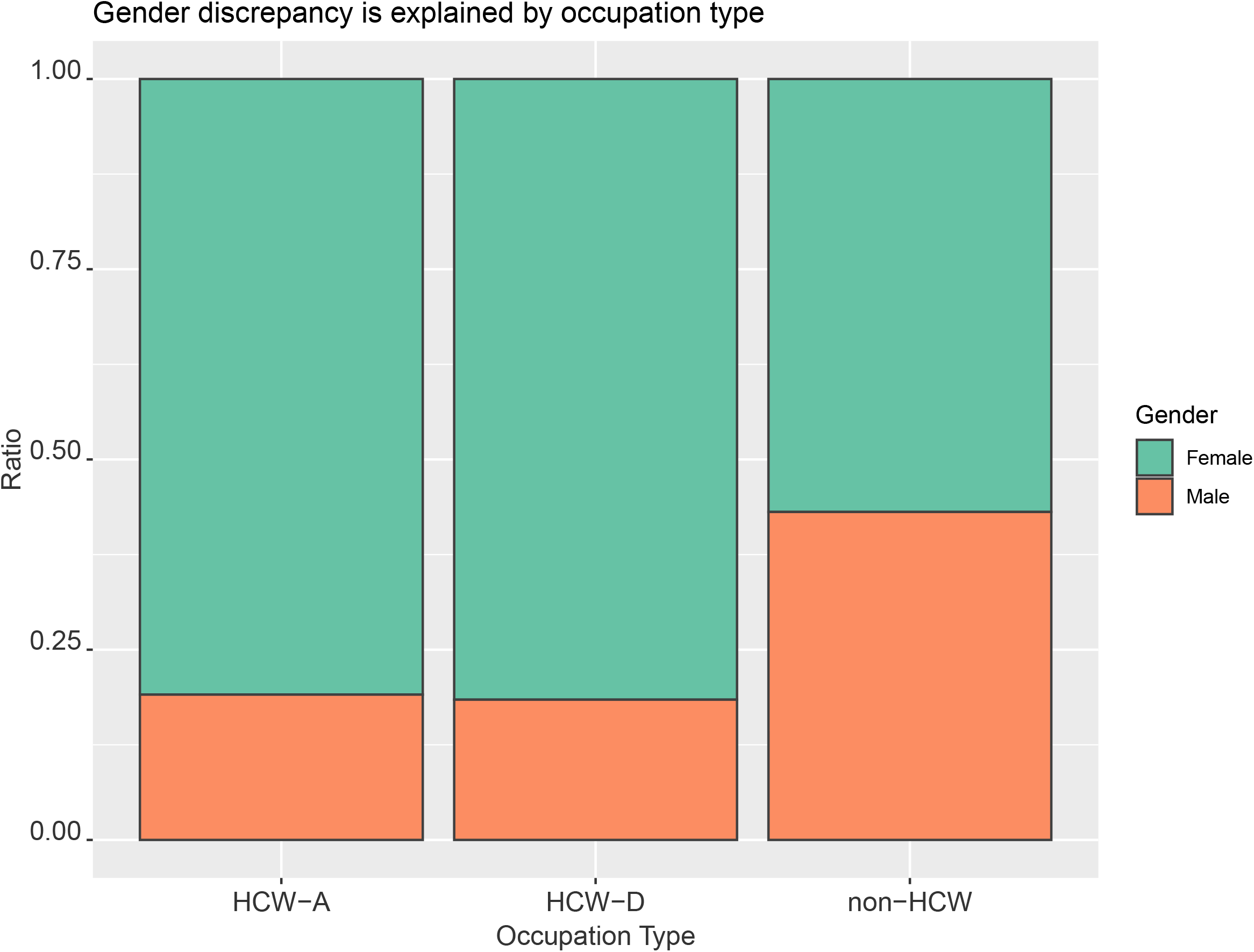
Gender discrepancy is explained by occupation type; 82% of HCW with direct patient contact (HCW-D) and 81% with indirect contact (HCW-A) versus 57% of other occupations were female in our data.

**Figure 2.**
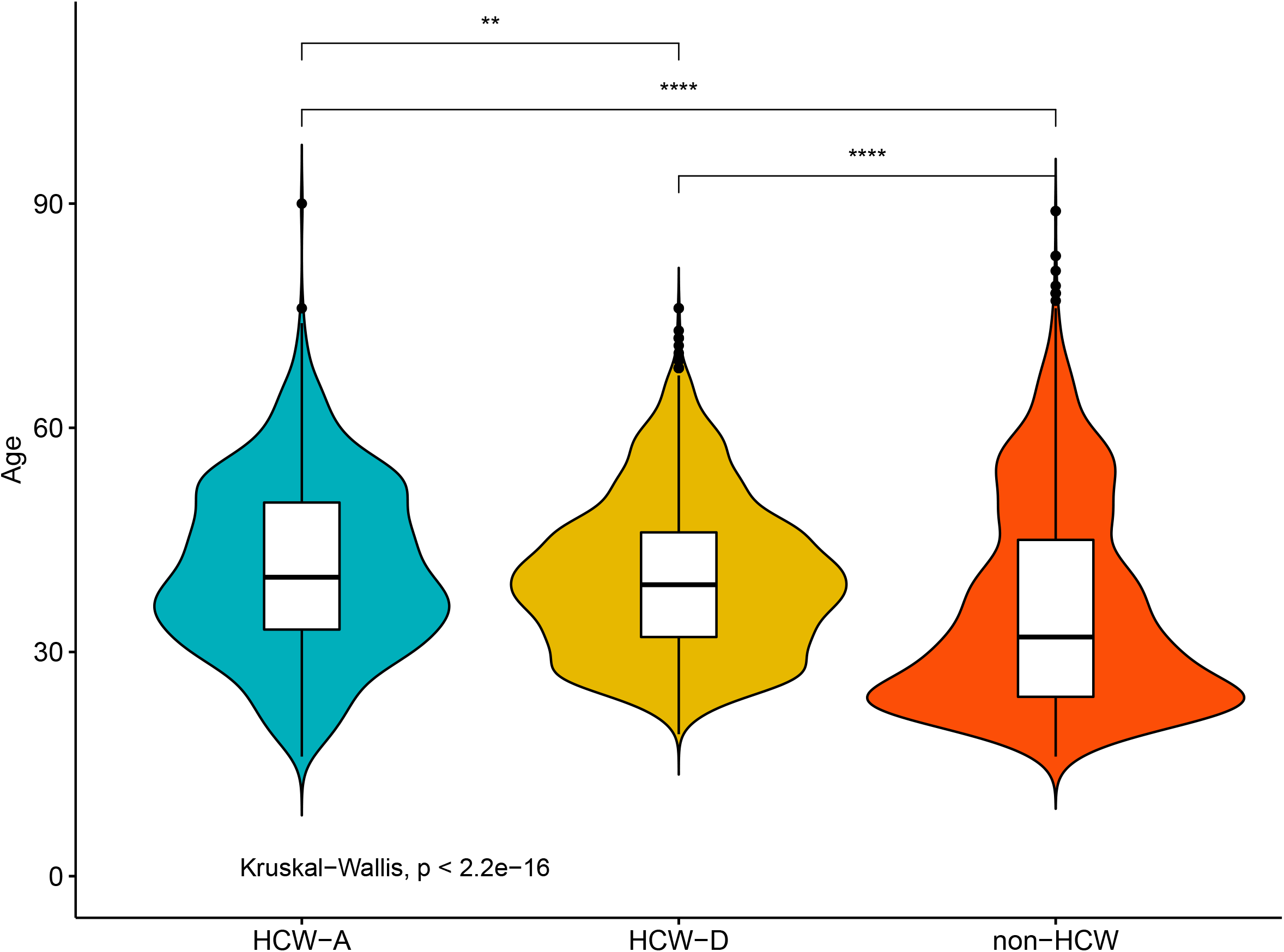
Age difference based on occupation type; 41.0 ±11.4 for HCW-D, 39.9 ±10.2 for HCW-A, and 35.6 ± 13.6 years for non-HCW. * p=0.004; **** p<0.0001.

We also combined both healthcare-related groups into one group (HCW) and compared them to non-HCW for easy interpretation of results. The odds ratio for positive COVID-19 testing was OR=1.5, 95% CI 1.35 – 1.67 (p<0.0001) for the HCW group versus non-HCW.

### Time analysis

Data was allocated into weekly time fractals. **Figure 3a** shows the number of positive and negative cases over the course of the data gathering. There were on average 137 tests performed per week with 32 of them returning positive (23.4%). The proportion of positive to negative cases slightly declined over time. The proportion of females to males significantly declined overtime the period (tau=-0.48, p<0.0001) (**Figure 4**).

**Figure 3.**
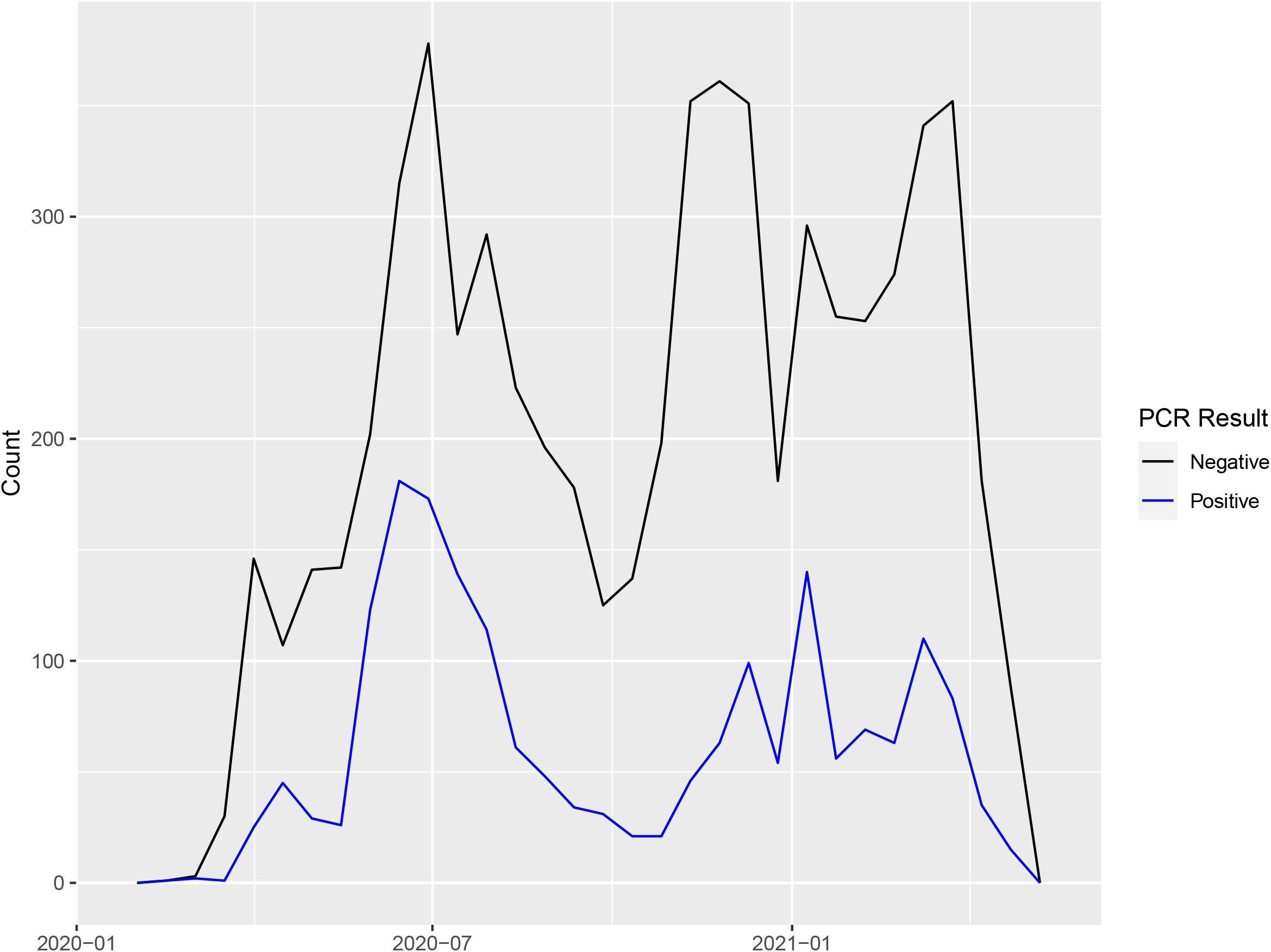
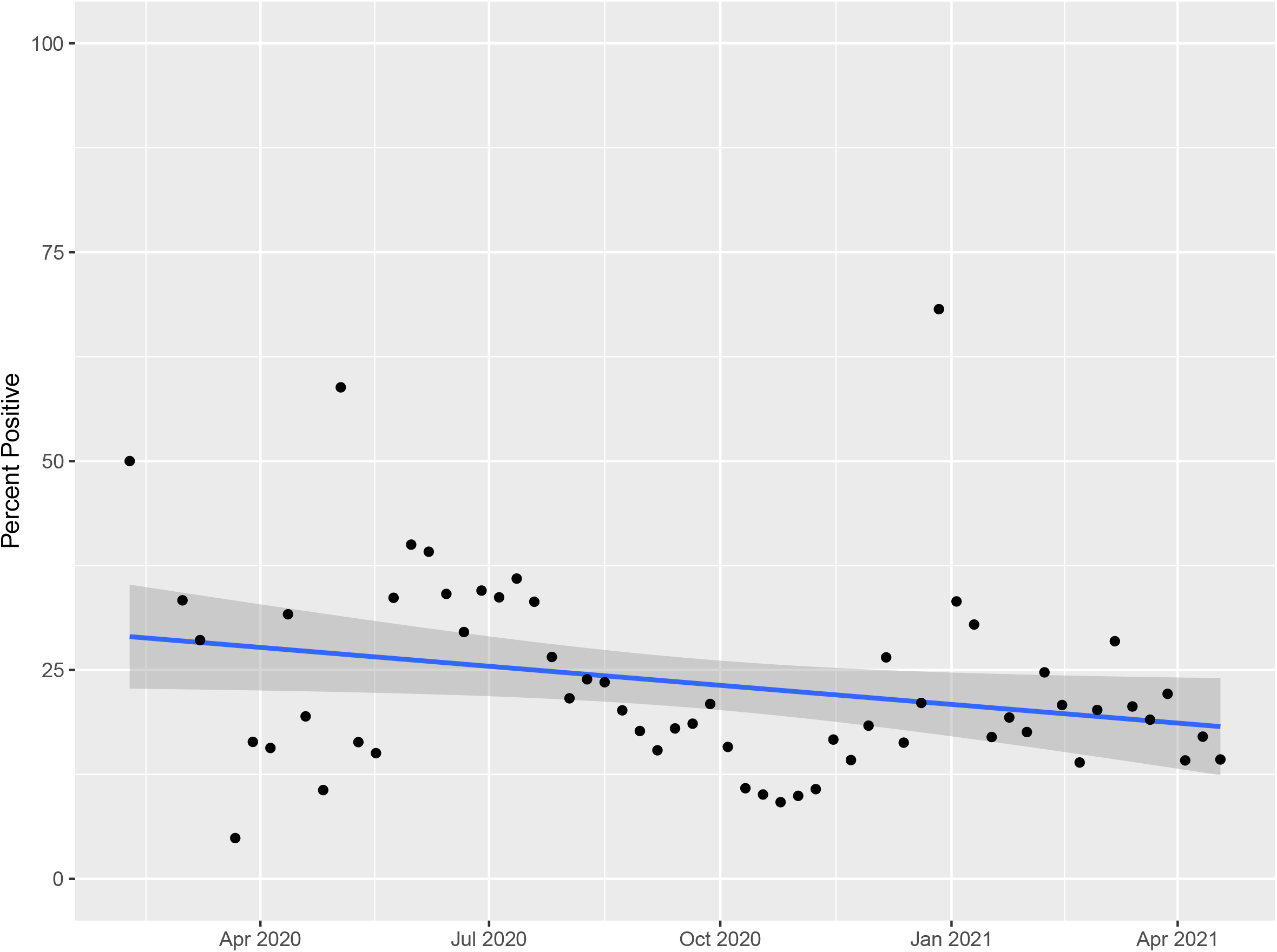
**a** Positive and negative test results over time. **b** Percent of positive results (compared to all tests) slightly declined over time (tau=-0.19, p=0.034).

**Figure 4.**
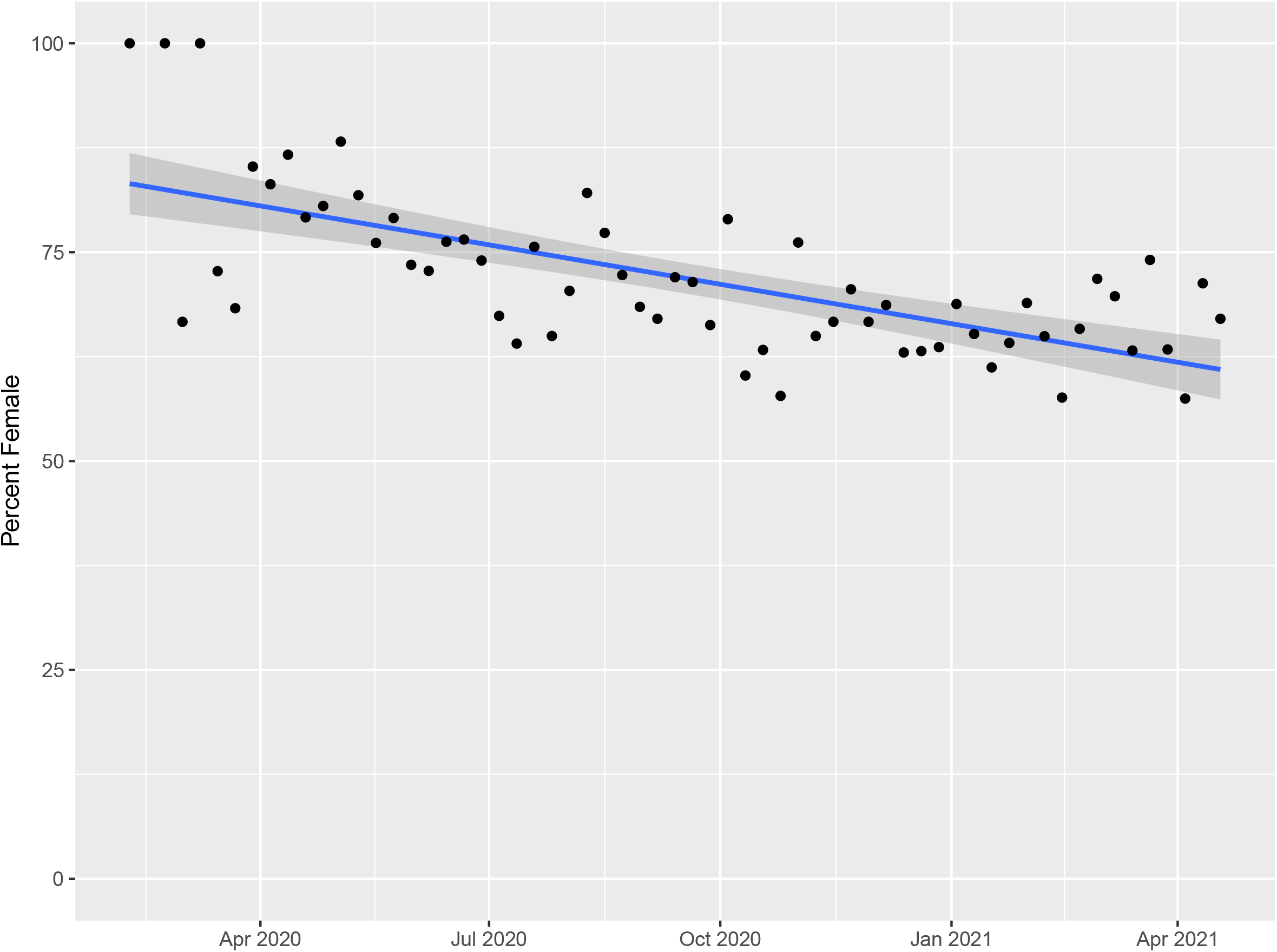
Percent of female patients declined over time (tau=-0.48, p<0.0001).

**Figure 5a** shows all tests taken over time based on job types. In the positive cases, we analyzed changes in the proportion of HCW (both direct and indirect contact) to non-HCW over time. At the beginning of the pandemic, HCW were the majority of the positive cases, however, their proportion declined significantly over the first 10 months (tau=-0.52, p<0.0001). In the final 4 months of our data gathering, the proportion of HCW remained steady (tau=+0.10, p = 0.62) (**Figure 5b**). Mann-Kendal trend test shows a steady decline in this ratio over the entire period (tau=-0.58, p<0.0001).

**Figure 5.**
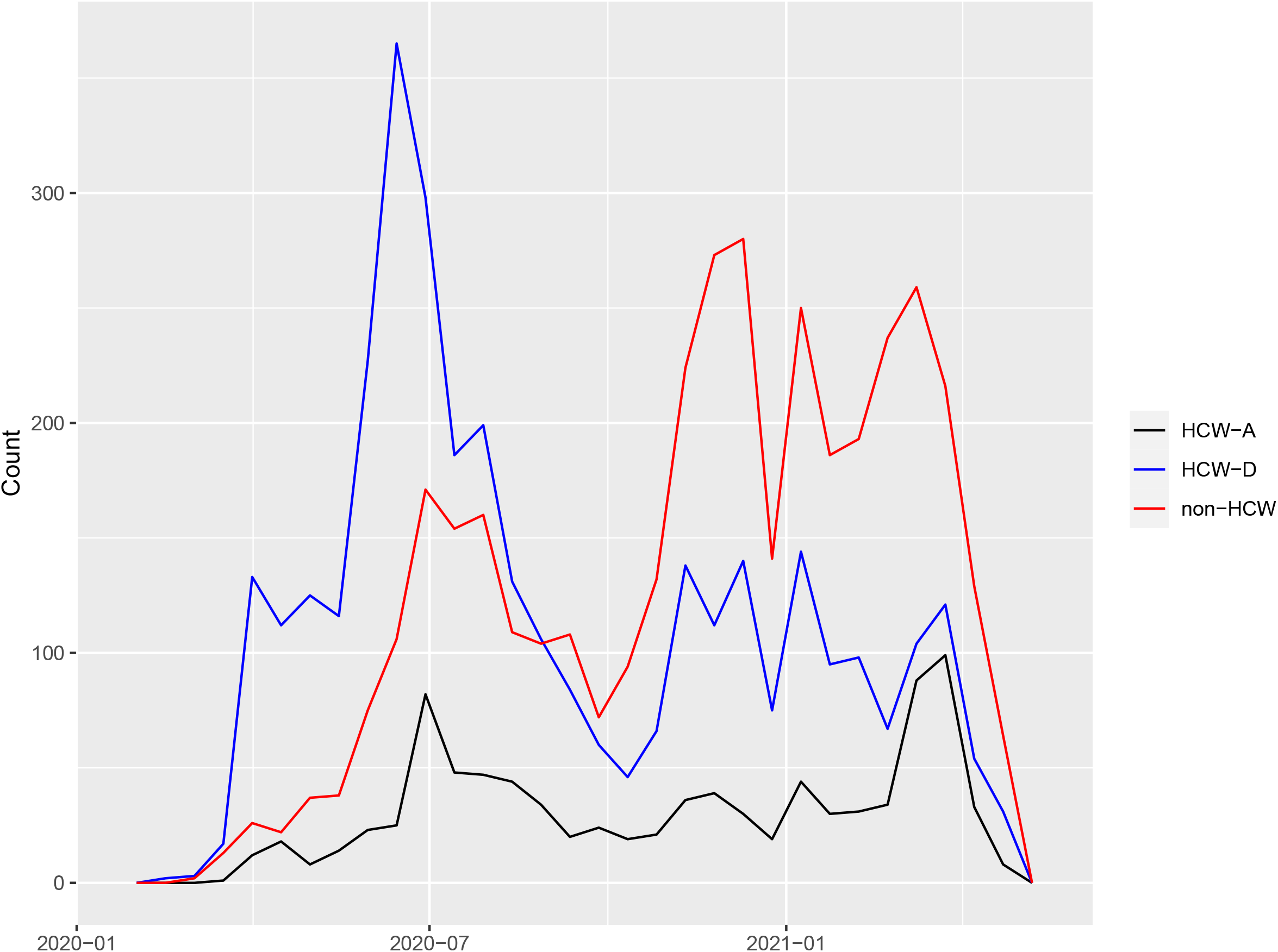
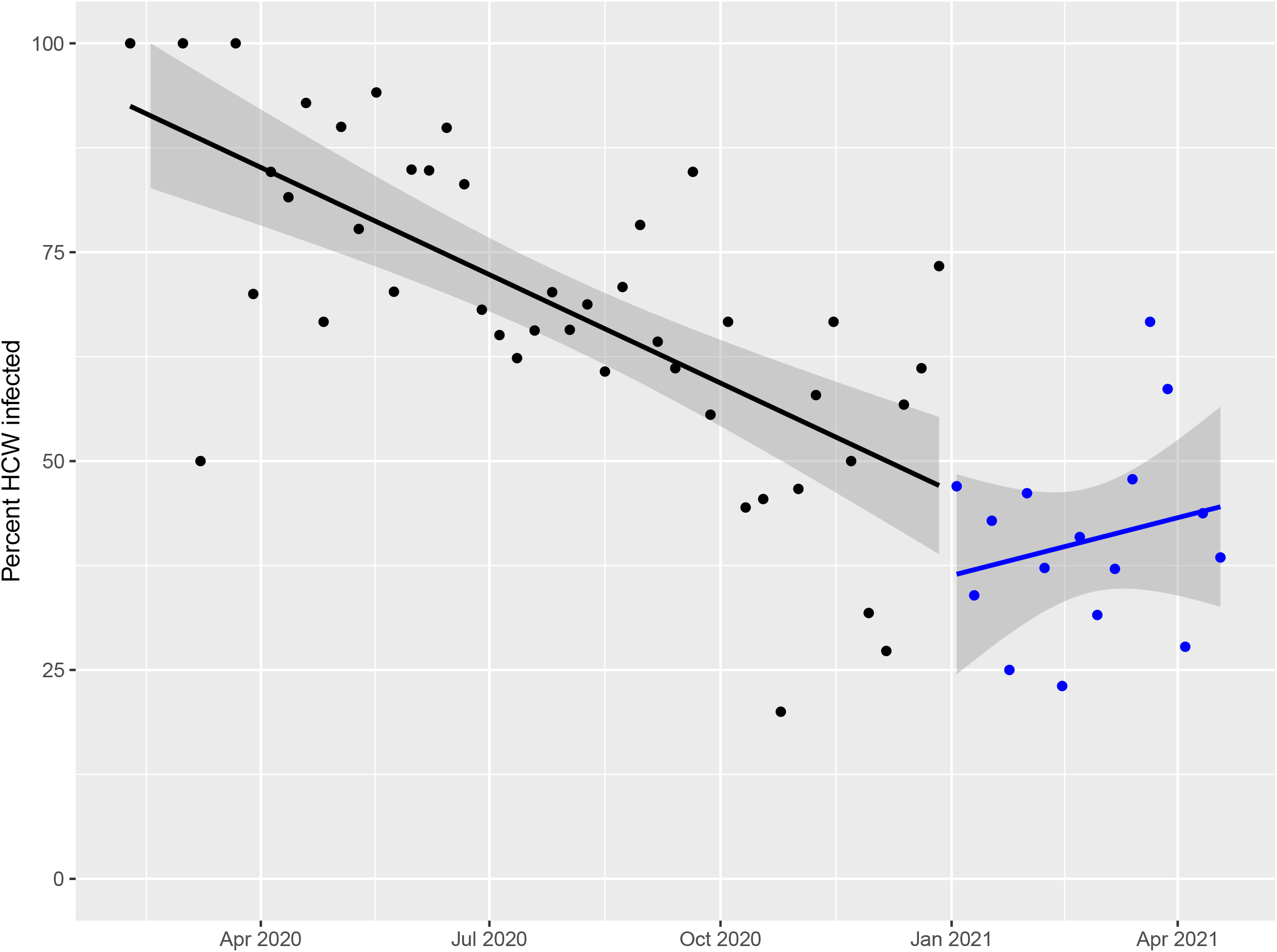
**a** Count of all patients based on occupation type. HCW predominance in the beginning months of the pandemic versus non-HCW in the final months of the data gathering. **b** COVID-19 positivity among HCW (both direct and indirect) compared to all positive patients declined rapidly (tau=-0.52, p<0.0001) in the first months of the pandemic, then it stays relatively steady over the rest of the data gathering (tau =+0.10, p=0.62).

### Address heat map

A digital map of all patient zip codes was created using Leaflet JavaScript library maps and it is available online at http://wdchealth.covid-map.com/shiny/covid-map/. Page one of this website shows a heat map of all positive cases. Page two shows a time series of individual cases based on their zip codes. In the time series, each dot represents a single new case and it stays on the map for 14 days. Cut scenes of the time series are shown on the Temporal Map Analysis page. The “Count of Cases by Zip Code” table on the website shows the top ten zip codes with the highest number of positive cases.

### Forecast model

We calculated the Hurst parameter for positive and negative cases, yielding 0.78 and 0.84, respectively. It revealed the self-similarity of this infection and a pattern that persists with a long-memory process of over seven months, **Figure 6**. The Autocorrelation Function (ACF) plot indicates that the memory indeed persists over the range of data collection once exposed in a community. Negative cases also showed a long-memory process as the long memory process crosses the axis on the autocorrelation function with persistent memory. We used our model on 1,914 positive tests, aggregating them to the weekly counts to estimate future weekly new cases

**Figure 6.**
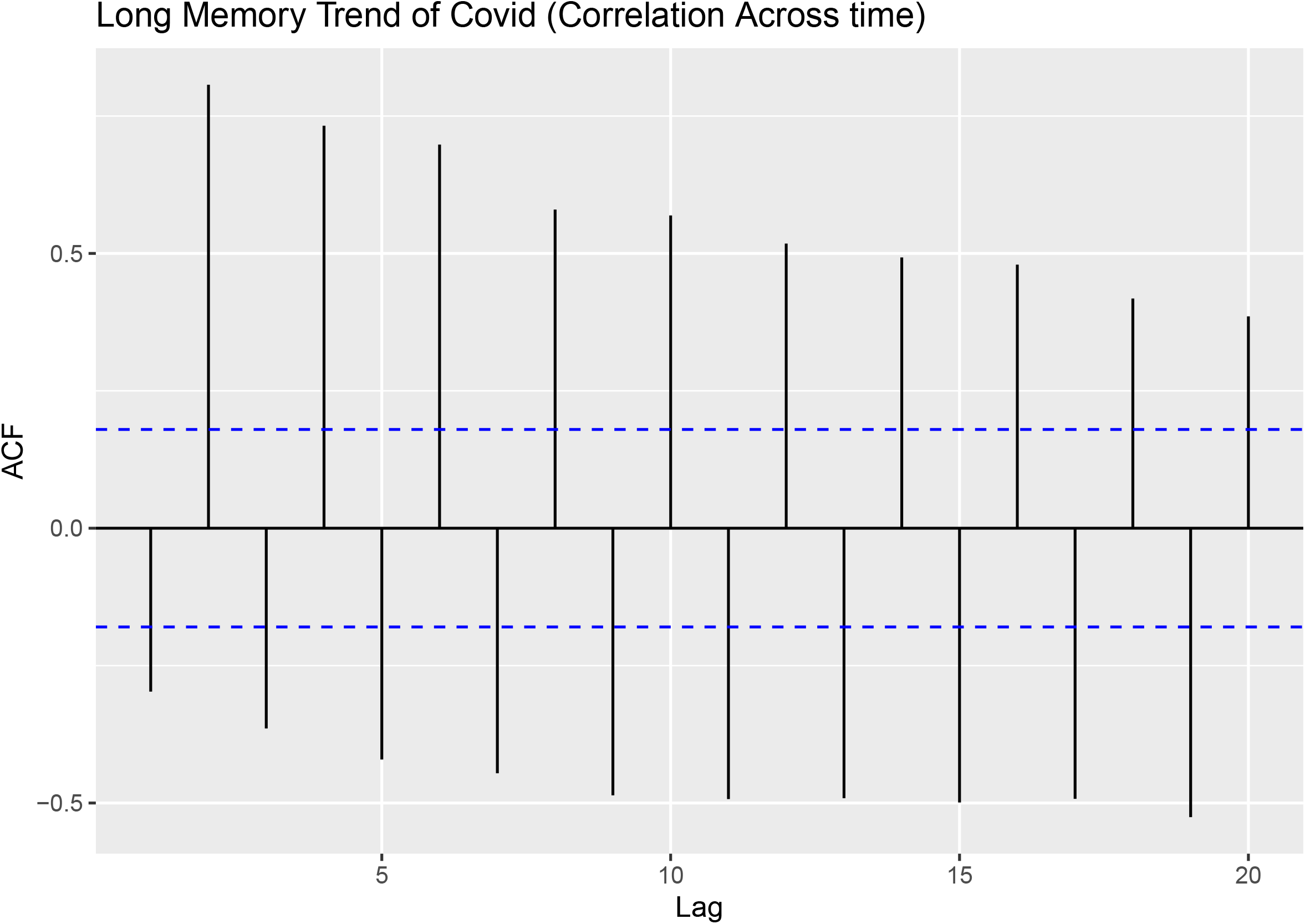
Auto Correlation Function (ACF) plot shows a long memory process of both positive and negative cases.

(**Figure 7**). It shows a steady slow increase of future cases (purple region) of infection. The purple hues are the statistical bounds (80% and 95%) of our prediction, while the dark blue line indicates the prediction of future cases. **Figure 8** shows the “fractal dimension plot” and the long memory process of the virus.

**Figure 7.**
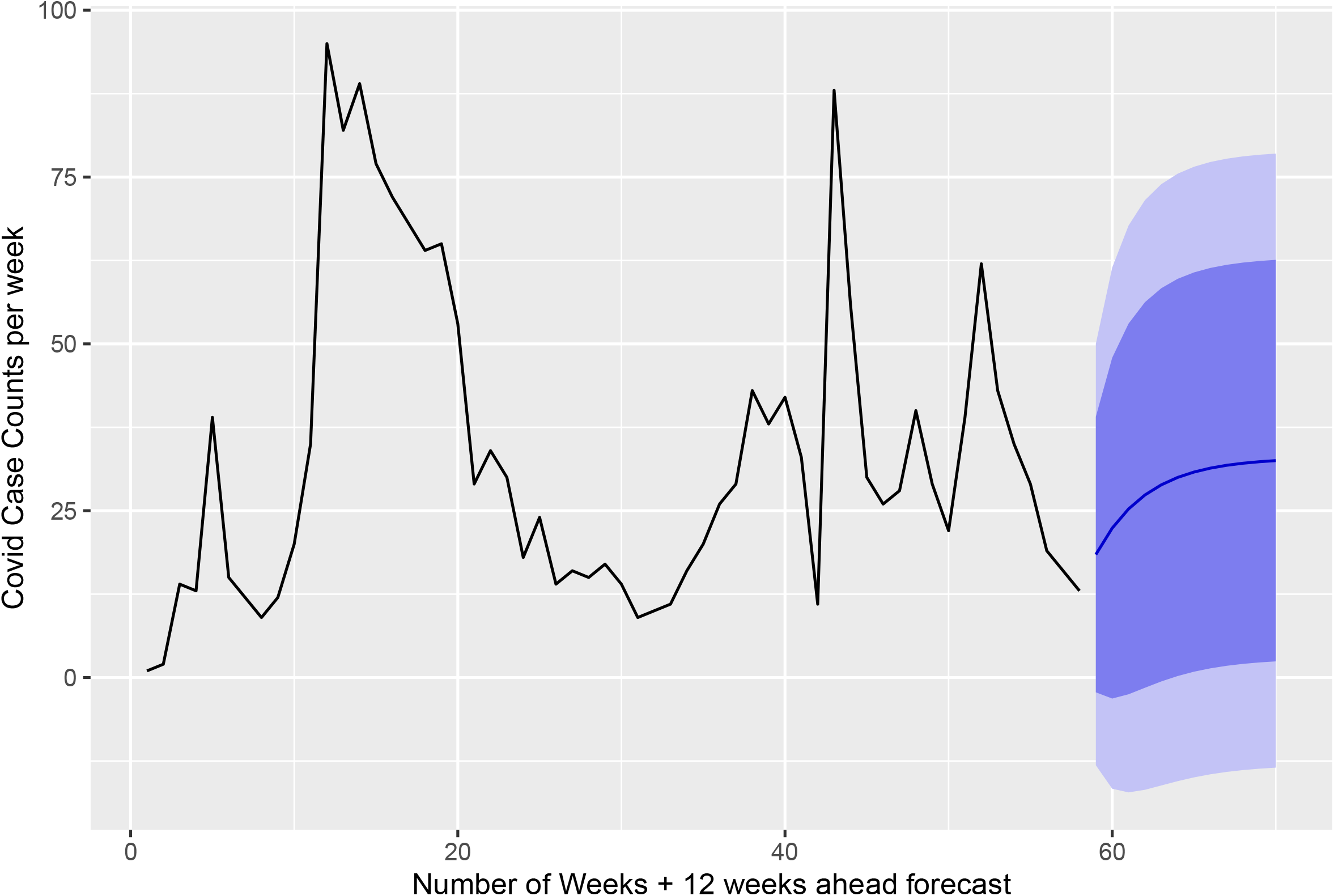
Count of all positive cases over the course of our data gathering and the ARFIMA prediction of the final 12 weeks.

**Figure 8.**
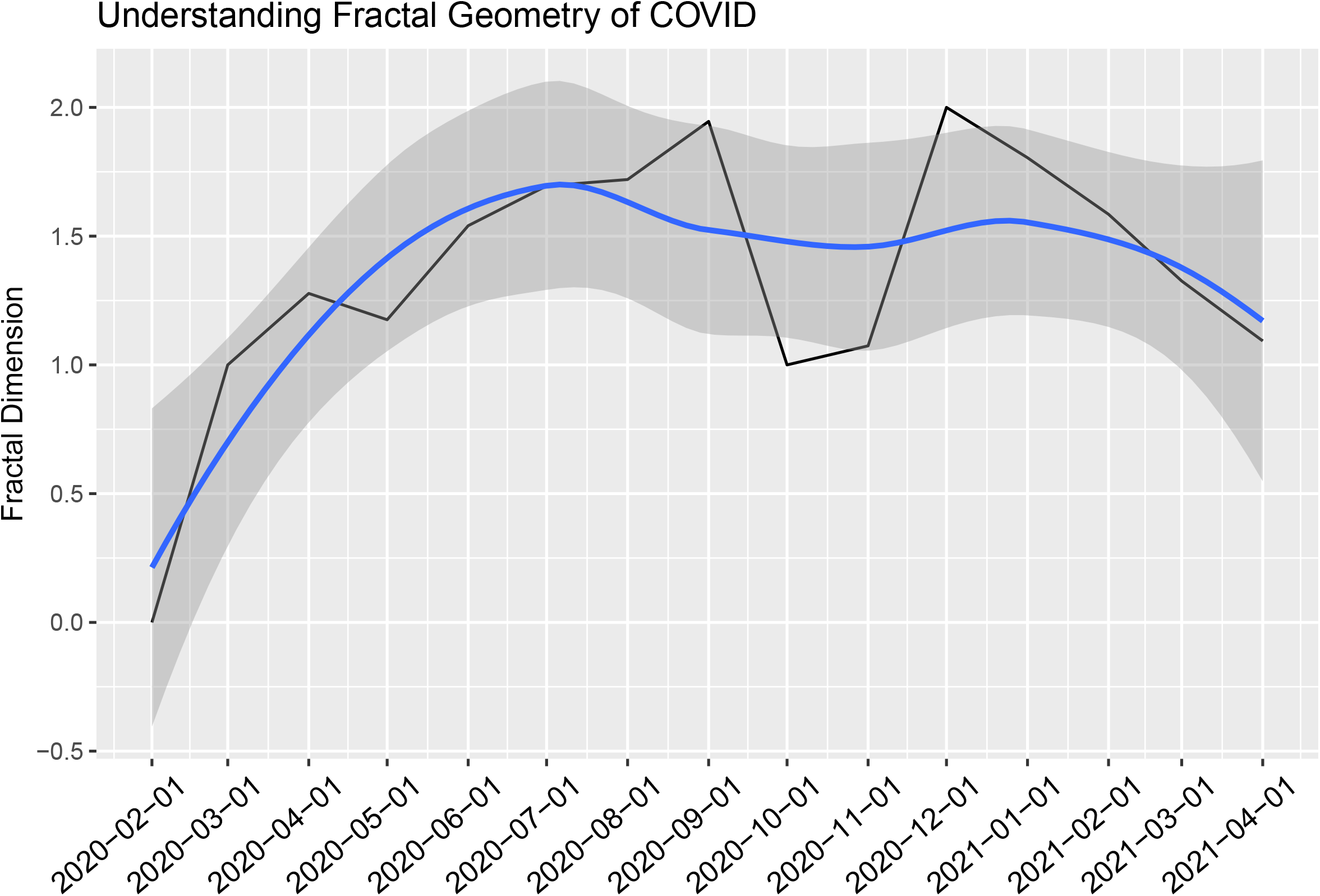
The fractal dimensions plot shows a long memory process (6-7 months) for COVID-19. The black line is the actual fractal dimension and the blue line is the LOESS smoothing of the black line of positive cases for each month.

### Testing the model

The ARFIMA prediction model was created using data available until April 2021. In July 2021 we gathered data of the past 3 months (“actual value”) and compared it to the “predicted values”. **Figure 9** compares predicted (red line) versus actual (blue lines) count of positive cases. The average error was 1.9 cases per week. We used this model to forecast the count of new positive cases in the next 12 weeks (August to October 2021). The new graph is available online at http://wdchealth.covid-map.com/shiny/covid-map/.

**Figure 9.**
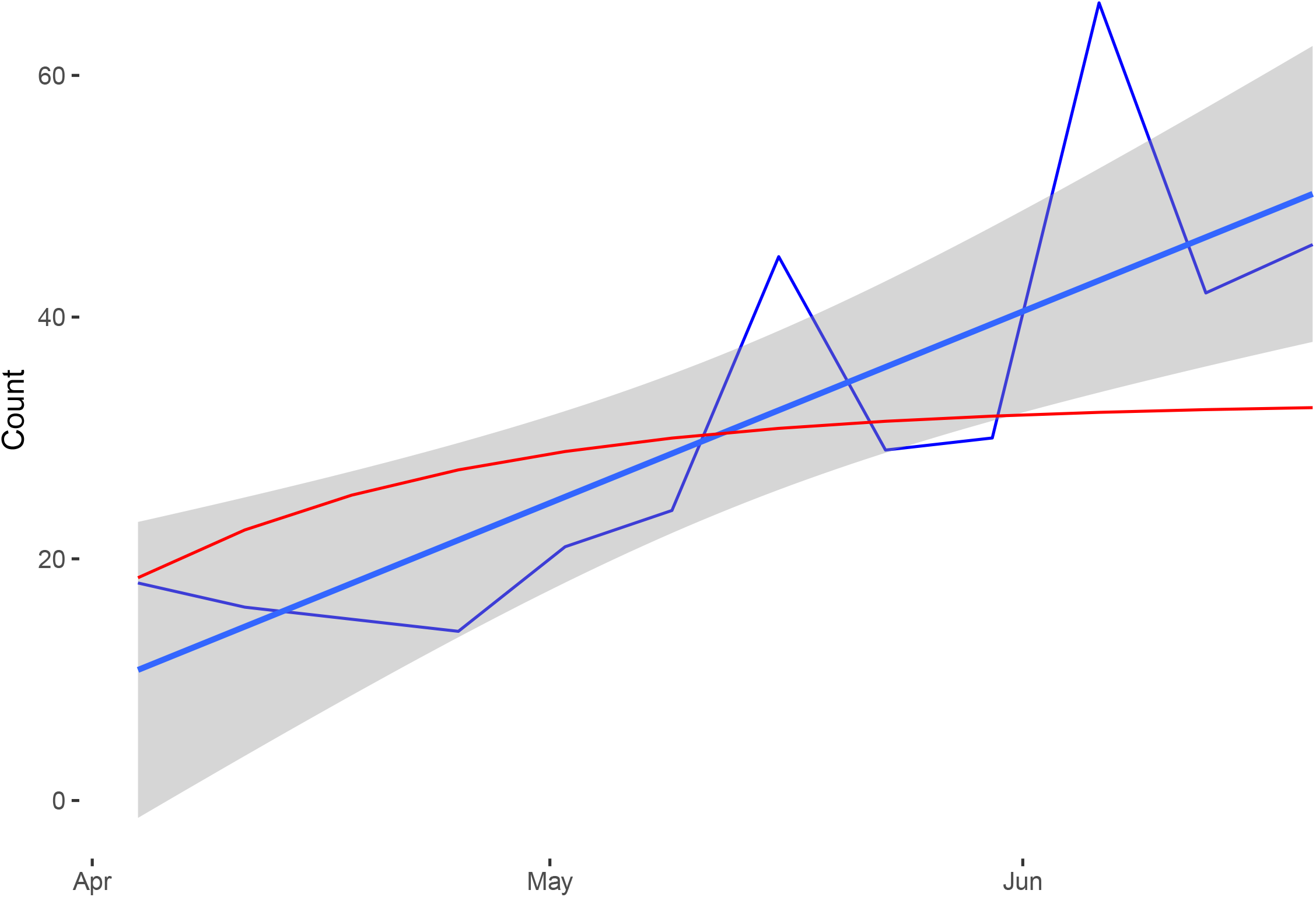
Testing our forecast model: Comparison of predicted count of positive cases (red line) versus count of actual cases (blue lines) in the past 3 months. The average error was 1.9 cases per week.

## DISCUSSION

This is one of the largest cohorts to date of COVID-19 tests in non-hospitalized populations. This data is crucial since the majority of these patients do not need medical assistance or hospitalization while most published literature regard hospitalized patients. Our outpatient data, collected over a relatively long time (14 months), bring useful information on transmission waves of COVID-19, and the effects of quarantines.

The covered area includes two large universities and multiple hospitals and clinics. Thus, the population included many healthcare workers (with both direct and indirect patient contact) and also young university students, which made it possible to compare the groups. We calculated the odds of infection in HCW to be 1.5-fold higher than non-HCW not regarding temporal changes.

Once we included time in the analysis, however, a new picture was revealed, showing the HCW to be the main bulk of the infected population at the beginning of the pandemic. The time analysis graph shows a rapid decline in the proportion of healthcare-related patients as the number of infected increased in the early stages of the pandemic. Once the infection spread to the general population, the proportion of healthcare-related patients remained steady. It is important to keep in mind that our data is from university medical center, which receive university employees as their main patients. However, in the area covered, there are no other COVID-19 testing locations.

Li et al. [13] brought the first report on the topic of healthcare workers in January 2020, showing 15 healthcare workers infection in a population of 370 patients, corresponding to 4.05%. Then, Wang et al. [14] reported 28.9% in a total 138 population, and Feng [15] reported a large study with 44,672 patients with a proportion of 3.84% of HCW. A meta-analysis with data until April-23, 2020, showed that HCW were 4.2% of positive tests in China, 17.8% in the USA, and 9% in Italy [5]. Considering the HCW population, the first screening study, in England, found positive tests in 5% on March 10-11, 2020, and growth to 20% on March 30-31 [16]. Besides the high proportion, it shows the high doubling time during a period of exponential growth. Eyre et al. showed that in-hospital function and place of work also influence the risk, they found that HCW positivity varied from 28% in acute medicine, 12% in the emergency department, and only 9.8% in intensive care units [17]. That may be related to many aspects ranging from treated patients’ diagnosis to the availability and knowledge of protection equipment usage.

Our study shows how HCW were exposed during the first year of the pandemic and how it increased the risk of SARS-CoV-2 infection. The high percentage of healthcare workers and the university environment can explain the mean age at the tested population, which was only 38 years old, and it is similar to other reports about HCW [17-18]. It also explains the gender discrepancy: the female gender represented 82% of HCW-D and 81% of HCW-A, which agrees with the 2019 WHO report on gender discrepancy in HCW. Furthermore, women are more likely to seek medical care in case of non-urgent ailments, which can also be a cultural aspect [19]. The gender discrepancy requires caution at the interpretation of the results, but it was not related to COVID positivity (OR=1.12, p=0.041). Mann-Kendall analysis showed that the female proportion slightly decreased over time, which could relate to the spread of the disease from healthcare workers to the general population.

It is important to notice that vaccination started at the end of January 2021 at a low-speed pace in the studied population. Thus, about 85% of the evaluated time was before vaccination. Moreover, the data reflects a period before the arrival of COVID-19 variants. Indeed, the first detected in Brazil was the P1 variant and it was very related to the healthcare system in the northern region of Brazil. Those variants were found near our city only on February 21, so it did not impact our data.

The test positivity evaluated along time showed SARS-CoV-2 incidence oscillation along the first pandemic year (**Figure 3a**). As the fractal analysis shows, COVID-19 has a long-memory process; indicating that infection waves have residual effects for up to 6 to 7 months. That information is important to build health policies that should take into count a long period, at least 7 months.

The city of Campinas had quarantine during the first two months (April and May), then activities were reopened progressively and oscillated from 30 to 80%. During the periods of most restriction, people’s circulation ranged at about 40%. Most of the regular business restarted after August, including restaurants, bars, and malls, with restrictions on people and working hours. As one can see in **Figure 3a**, during those months of business reopening, the number of cases had decreased. However, based on our forecast analysis, COVID-19 has a long memory process, meaning a short period of quarantine lift is unlikely to have a major effect on the surging of new cases.

### Fractal dimensions and forecast

The existence of a long memory process was first explored by Hurst (1951) as a solution for regulating the flow of the Nile River. The Nile River example has since become the most famous example of the existence of a long memory process: long periods of high flow levels were followed by long periods of low flow levels as Hurst observed. Mandelbrot and van Ness (1968) introduced the Hurst parameter (H) to describe the long-term memory of a time-series process. As H gets closer to 1, the more persistent the time series is considered and at values less than or equal to 0.5, the long-memory process does not exist. It is important to note that the Hurst parameter can only be approximated. We used Whittle’s approximation using the maximum likelihood estimation (MLE) approach to estimate the Hurst parameter [12].

In our study, the “fractal dimension plot” (**Figure 8**) shows that while quarantines might help with reducing the spread of the virus, it does not stop the spread due to its long memory process. **Figure 8** shows that once the infection has begun, it is not possible to slow it down until the virus has run through its infection period but also puts other members of a community at risk, given its statistical self-similarity and fractal nature. This implication can be readily observed in the HCW community. An interesting aspect of this analysis is the long-memory process of negative cases as we can observe in **Figure 6**. This shows that while the virus remains effective in infections over time, once the population adapts to this reality, the infection rate slows down considerably and as our plot shows, we are still, and persistently, in a slowdown phase of the infection period.

Comparing predicted values to actual values three months later (May-July 2021) showed that our model is a reliable tool to forecast new cases with an average error of ±1.9 cases. A model that can predict future new infection with an error of only ±2 patients per week can be a very valuable tool in policy making and healthcare budgeting. Our ARFIMA model used the fractal nature of new COVID-19 cases and counted the long memory process to create a more reliable forecast tool. There are already many published predicting tools to forecast new COVID-19 cases and many of them have shown poor reliability. Not counting the long memory process of this virus can explain the poor reliability of the already published models of this disease. Another reason could be these models were created very early in the course of the pandemic using data gathered in very short amount of time.

Our study is limited by its population, which is younger and with a higher female proportion than the general population and other studies populations due to healthcare representativeness, reaching about half of the study population. Also, it brings evidence from a period before vaccines and before COVID-19 variants, thus it cannot access the impact of neither one. However, it made possible to further understand the pandemic dynamics among HCW with direct or indirect patient contact versus non-HCW. It brings important evidence of the pandemic behavior which may be helpful for planning public policies and prepare HCW and providers for current and future epidemic diseases in our community.

## CONCLUSION

Health care workers and associates have a sustained 50% higher risk of COVID-19 positivity in the pandemic. Time-series analysis showed that the virus spread mainly among HCWs in the first 6 months of the pandemic before it further spread into the general population. The SARS-CoV-2 virus has a long-memory infection pattern that persists over time lasting for months. We plan to routinely publish our prediction of the next 12 weeks on our website http://wdchealth.covid-map.com/shiny/covid-map/ and update it according to population previous infection, virus variants and vaccination impact.

## Data Availability

The data that support the findings of this study are available from the corresponding author upon reasonable request.

http://wdchealth.covid-map.com/shiny/covid-map/

## Author Contributions

MJ, LMG, and AAE: data analysis and manuscript writing.

PAFL, FAVD, KB, LSBDC, CFG, KLF, ACP, CLM: data collection and manuscript editing LOR: funding acquisition, project development, supervision.

## Acknowledgment

To the involved institution(s), the patients, and those that provided and cared for study patients.

## Research Involving Human Participants

University of Campinas ethics committee approval number: 4.173.069

## Data Availability Statement

**Supplement:** Heatmap cut scenes show the spread of the virus in the area covered over time.

## REFERENCES

1- Wu D, Wu T, Liu Q, Yang Z. The SARS-CoV-2 outbreak: What we know. Int J Infect Dis. 2020;94:44–48. doi:10.1016/j.ijid.2020.03.004.

2- Wang C, Horby PW, Hayden FG, Gao GF. A novel coronavirus outbreak of global health concern [published correction appears in Lancet. 2020 Jan 29]. Lancet. 2020;395(10223):470–473. doi:10.1016/S0140-6736(20)30185-9.

3- Prezioso C, Marcocci ME, Palamara AT, De Chiara G, Pietropaolo V. The “Three Italy” of the COVID-19 epidemic and the possible involvement of SARS-CoV-2 in triggering complications other than pneumonia. J Neurovirol. 2020;26(3):311–323. doi:10.1007/s13365-020-00862-z.

4- Rodríguez BO, Sánchez TL. The Psychosocial Impact of COVID-19 on health care workers. Int Braz J Urol. 2020;46(Suppl.1):195-200. doi:10.1590/S1677-5538.IBJU.2020.S124.

5- Sahu AK, Amrithanand VT, Mathew R, Aggarwal P, Nayer J, Bhoi S. COVID-19 in health care workers -A systematic review and meta-analysis. Am J Emerg Med. 2020;38(9):1727–1731. doi:10.1016/j.ajem.2020.05.113.

6- Nyashanu M, Pfende F, Ekpenyong M. Exploring the challenges faced by frontline workers in health and social care amid the COVID-19 pandemic: experiences of frontline workers in the English Midlands region, UK. J Interprof Care. 2020;34(5):655–661. doi:10.1080/13561820.2020.1792425.

7- Ming X, Ray C, Bandari M. Beyond the PPE shortage: Improperly fitting personal protective equipment and COVID-19 transmission among health care professionals. Hosp Pract (1995). 2020;48(5):246–247. doi:10.1080/21548331.2020.1802172.

8- Al-Hity S, Bhamra N, Kumar R, et al. Personal protective equipment guidance during a global pandemic: A statistical analysis of National perceived confidence, knowledge and educational deficits amongst UK-based doctors. Int J Clin Pract. 2021;75(5):e14029. doi:10.1111/ijcp.14029.

9- Nguyen LH, Drew DA, Joshi AD, et al. Risk of COVID-19 among frontline healthcare workers and the general community: a prospective cohort study. Preprint. medRxiv. 2020;2020.04.29.20084111. Published 2020 May 25. doi:10.1101/2020.04.29.20084111.

10- Bandyopadhyay S, Baticulon RE, Kadhum M, et al. Infection and mortality of healthcare workers worldwide from COVID-19: a systematic review. BMJ Glob Health. 2020;5(12):e003097. doi:10.1136/bmjgh-2020-003097.

11- Mendonça-Galaio L, Sacadura-Leite E, Raposo J, França D, Correia A, Lobo R, Soares J, Almeida C, Shapovalova O, Serranheira F, Sousa-Uva A. The COVID-19 Impact in Hospital Healthcare Workers: Development of an Occupational Health Risk Management Program. Port J Public Health 2020;38(Suppl 1):26–31. doi: 10.1159/000515327.

12- Beran, J., 1994. Statistics for Long-Memory Process, Boca Raton: CRC Press LLC.

13- Li Q, Guan X, Wu P, et al. Early Transmission Dynamics in Wuhan, China, of Novel Coronavirus-Infected Pneumonia. N Engl J Med. 2020;382(13):1199–1207. doi:10.1056/NEJMoa2001316.

14- Wang D, Hu B, Hu C, et al. Clinical Characteristics of 138 Hospitalized Patients With 2019 Novel Coronavirus-Infected Pneumonia in Wuhan, China [published correction appears in JAMA. 2021 Mar 16;325(11):1113]. JAMA. 2020;323(11):1061–1069. doi:10.1001/jama.2020.1585.

15- Epidemiology Working Group for NCIP Epidemic Response, Chinese Center for Disease Control and Prevention. Zhonghua Liu Xing Bing Xue Za Zhi. 2020;41(2):145–151. doi:10.3760/cma.j.issn.0254-6450.2020.02.003.

16- Hunter E, Price DA, Murphy E, et al. First experience of COVID-19 screening of health-care workers in England. Lancet. 2020;395(10234):e77–e78. doi:10.1016/S0140-6736(20)30970-3

17- Eyre DW, Lumley SF, O’Donnell D, et al. Differential occupational risks to healthcare workers from SARS-CoV-2 observed during a prospective observational study. Elife. 2020;9:e60675. Published 2020 Aug 21. doi:10.7554/eLife.60675.

18- Mandana Gholami, Iman Fawad, Sidra Shadan, Rashed Rowaiee, HedaietAllah Ghanem, Amar Hassan Khamis, Samuel B. Ho. COVID-19 and healthcare workers: A systematic review and meta-analysis. International Journal of Infectious Diseases. 2021;(104): 335–346. Doi: 10.1016/j.ijid.2021.01.013.

19- Couto MT, Pineiro TF, Valença AO, Machin R, Silva GSN, Gomes R, et al. O homem na atenção primária à saúde: discutindo (in)visibilidade a partir da perspectiva de gênero. Interface Comum Saúde Educ, 2010, p. 257–270.

